# Sustained high prevalence of COVID-19 deaths from a systematic post-mortem study in Lusaka, Zambia: one year later

**DOI:** 10.1101/2022.03.08.22272087

**Authors:** Christopher J Gill, Lawrence Mwananyanda, William MacLeod, Geoffrey Kwenda, Rachel Pieciak, Lauren Etter, Daniel J. Bridges, Chilufya Chikoti, Sarah Chirwa, Charles Chimoga, Leah Forman, Ben Katowa, Rotem Lapidot, James Lungu, Japhet Matoba, Gift Mwinga, Benjamin Mubemba, Zachariah Mupila, Walter Muleya, Mulenga Mwenda, Benard Ngoma, Ruth Nkazwe, Diana Nzara, Natalie Pawlak, Lillian Pemba, Ngonda Saasa, Edgar Simulundu, Baron Yankonde, Donald M. Thea

## Abstract

**Background:** Sparse data documenting the impact of COVID-19 in Africa has fostered the belief that COVID-19 ‘skipped Africa’. We previously published results from a systematic postmortem surveillance at a busy inner-city morgue in Lusaka, Zambia. Between June-October 2020, we detected COVID-19 in 15-19% of all deaths and concentrated in community settings where testing for COVID-19 was absent. Yet these conclusions rested on a small cohort of 70 COVID-19+ decedents. Subsequently, we conducted a longer and far larger follow-on survey using and expanding on the same methodology.

**Methods:** We obtained a nasopharyngeal swab from each enrolled decedent and tested these using reverse transcriptase quantitative PCR (RT-qPCR). A subset of samples with a PCR cycle threshold <30 underwent genotyping to identify viral lineages. We weighted our results to adjust for enrolment ratios and stratified them by setting (facility vs. community), time of year, age, and location.

**Results:** From 1,118 enrolled decedents, COVID-19 was detected among 32.0% (358/1,116). We observed three waves of transmission that peaked in July 2020, January 2021, and ∼June 2021 (end of surveillance). These were dominated by the AE.1 lineage and the Beta and Delta variants, respectively. During peak transmission, COVID-19 was detected in ∼90% of all deaths. Roughly four COVID-19 deaths occurred in the community for every facility death. Antemortem testing occurred for 52.6% (302/574) of facility deaths but only 1.8% (10/544) of community deaths and overall, only ∼10% of COVID-19+ deaths were identified in life.

**Conclusions:** COVID-19 had a devastating impact in Lusaka. COVID-19+ deaths occurred in all age groups and was the leading cause of death during peak transmission periods. Testing was rarely done for the vast majority of COVID-19 deaths that occurred in the community, yielding a substantial undercount.

**What is already known on this topic:**

- Previously, we reported that COVID-19 was present among 15-19% of all decedents passing through a busy city morgue in Lusaka.
- Data documenting the mortal impact of COVID-19 in Africa remain sparse.
- Several modeling groups have also argued that COVID-19’s impact in Africa has been underreported and hence underestimated.

**What this study adds:**

- Antemortem testing for COVID-19 captured only ∼10% of COVID-19 positive individuals indicating a substantial gap in surveillance.
- During peak transmission periods, ∼90% of all deceased individuals tested positive for COVID-19.
- Most COVID-19 positive deceased adults presented with symptoms typical of COVID-19, arguing that COVID-19 caused their deaths and was not a co-incidental finding.
- Deaths occurred across the age spectrum, including among young children, indicating a different pattern of impact from what has been seen in high income country settings.
- We document three waves of transmission, attributable to the AE.1 lineage, and the Beta and Delta variants, respectively.

## INTRODUCTION

Despite the evident harm from the COVID-19 pandemic globally, accurate data about the pandemic’s mortal impact in Africa remain sparse. In early 2021, our team reported results from systematic COVID-19 post-mortem surveillance conducted from June to October 2020 at the central morgue in Lusaka, Zambia.^1^ Using PCR, we detected COVID-19 in nearly 20% of decedents, most of whom had died in the community. In that group, none had been tested for COVID-19 prior to death. Yet, even among facility deaths antemortem testing was uncommon. Hence, COVID-19 deaths only appeared rare in Lusaka because testing was rarely done. We theorized that the so-called ‘Africa Paradox’^2-7^ was a myth born from insufficient surveillance data. Other groups have reached similar conclusions.^8 9^

Our analysis yielded other provocative findings. In contrast with the US and other high-income country (HIC) settings where deaths are concentrated in the elderly, COVID-19+ deaths in Lusaka occurred more evenly across the age spectrum. Further, 10% of COVID-19+ deaths occurred in children, a surprising finding given how rare pediatric COVID-19 deaths are in HICs. It remains an open question whether these pediatric deaths were caused by COVID-19 or whether the virus was a coincidental finding.

These conclusions were conservative given that our analysis rested on only seventy COVID-19+ decedents. Subsequently, our team conducted a second round of post-mortem COVID-19 surveillance, spanning the period January through June 2021. In this larger study we again quantified the burden of fatal COVID-19 by setting (community vs. facility deaths), calendar date, and geography; and observed the impact and patterns of clinical presentation among children vs. adults. In addition, we aligned our prevalence data with genetic sequencing to characterize shifts in viral lineages/variants over time. Consistent with our prior report, COVID-19 had a severe impact in Lusaka. Across both surveillance periods, we document three waves of transmission.

## METHODS

### Overview

Our COVID-19 surveillance work builds upon a larger body of postmortem surveillance work initiated to define the fatal burden of Respiratory Syncytial Virus and *Bordetella pertussis*. With the pandemic, we amended our protocols to expand surveillance to all age ranges and to test for COVID-19. The RSV results have now been published,^10-12^ and the pertussis results are currently under review.

We direct the readers to our prior publication for complete details of the surveillance methods.^1^ Throughout this paper, we refer to several rounds of surveillance. Round 1 ran from June-Oct 2020; Round 2, comprising the new data in this paper, ran from Jan-June 2021. Round 3 is ongoing, was initiated early in 2022, and is planned to run through January 2023.

The research was conducted with the approval of the ethical review boards from Boston University and the University of Zambia; written informed consent was obtained from the next of kin or family representative in all cases.

### Sampling strategy

When capturing a high proportion of all deaths, postmortem surveillance is robust against ascertainment biases and provides a direct and unambiguous measure of a disease’s impact. We enrolled deceased individuals at the University Teaching Hospital (UTH) morgue. Given concerns about viral RNA degradation leading to false negative results, enrollment was restricted to deaths within the preceding 48 hours.

UTH is the main teaching hospital for the University of Zambia Medical School. Its morgue is the largest in the city and contributes/accounts for 80% of deaths of burial certificates issued by the council office in the city. This, plus the legal requirement to obtain a burial certificate, most of which are obtained from the UTH medical examiner’s office, makes it highly representative of all deaths in Lusaka.

Due to the high volume in the morgue and finite capacity of our team, we capped enrollments at ∼5-6 deaths per day. Also due to high volume, we enrolled community deaths at a 1:3 ratio, while enrolling the less common facility deaths at a 1:1 ratio. In response to the surprising high proportion of pediatric deaths in surveillance Round 1, we elected to also oversample this group enrolling infants (<1 year) at a 1:1 ratio. We have adjusted for these ratios in our total population prevalence estimates (see statistical analysis). Our team had no access to the clinical data about each decedent and could not know the PCR results prior to consent and enrollment. Therefore, we were confident that our enrolment strategy was robust against selection biases with respect to COVID-19 status.

Concurrently, we extracted total deaths by age stratum from the official government burial registry. This allowed us to estimate the proportion of all deaths represented by our sample, and, by comparing the age by death distribution of the two groups, assess whether the enrolled sample appeared broadly representative of the total.

### Data collection and case definitions

We defined a ‘facility death’ as one that occurred under care at UTH or a referring facility. We defined a community death as one that occurred outside of medical care. We defined pediatric deaths as those occurring between 0-≤19 years.

Following enrollment, individual and household demographic data were collected on all decedents, along with any antemortem COVID-19 testing results that were documented in the medical chart (for facility deaths) or reported by the next of kin/representative of the decedent during the verbal autopsy (for community deaths).

To infer the causal role of COVID-19 in deaths, we focused on the clinical data describing symptoms during the fatal event. For facility deaths, these data were obtained from a medical chart extraction focusing on symptoms at initial presentation to UTH; for community deaths, we conducted a verbal autopsy using the abbreviated tool validated by the Institute for Health Metrics Evaluation (IHME).^13^ From these syndromic data, we classified adult decedents (>19 years) into the following categories:

1. <u>Probable COVID-19 deaths</u>, where the individual presented with any combination of fever and/or respiratory symptoms;
2. <u>Possible COVID-19 deaths</u>, where the individual presented without classic respiratory symptoms, but rather with known sequalae of COVID-19 suggestive of a heart attack, stroke, or other acute vascular event related to COVID-19’s propensity to increase the risk of clotting disorders;^14^
3. <u>Probably not COVID-19 deaths</u>, where the individual had an exonerating cause of death suggesting that the COVID-19 finding on PCR was incidental.
4. <u>Unknown</u>, where the data were insufficient to adjudicate.

The reasons we only applied this strategy to adults is that it quickly proved ill-suited for children. Unlike adults, where fever and/or respiratory symptoms predominated, the pediatric presentations were more variable and respiratory symptoms less common. Since we were (and remain) uncertain about how COVID-19 should present clinically in African children, creating *a priori* case definitions based on how the virus behaves in adults could be misleading. Similarly, applying a new definition developed iteratively from the constellation of symptoms observed in children and then applying that definition forwards without validation against an external gold standard (such as histopathology coupled with PCR), constitutes a logical circularity.

Accepting this uncertainty, we opted simply to describe the pediatric case presentations without attempting to infer causality. The presenting symptoms largely clustered into two syndromic categories: (1) respiratory, which included any combination of upper and lower respiratory symptoms; and (2) gastro-intestinal, which included any combination of vomiting, diarrhea, and abdominal pain. Since fever could occur in either syndrome, or in isolation, we reported this separately. We then tallied these within different age groups from 0-19 years to observe if the symptomatology was age dependent.

### Biological sample collection

From each deceased individual, we obtained posterior nasopharyngeal (NP) samples. We used flocked-tipped NP swabs (Copan Diagnostics, Murietta, CA, USA),^15 16^ inserting the swab into both nares and then rotating 180 degrees in both directions and collected into 3mL vials of universal transport media. Samples were stored cold or on ice until processing at our onsite molecular lab.

### Laboratory procedures

#### PCR testing for COVID-19

After vortexing to elute samples, total nucleic acid for PCR was extracted using the EasyMAG system (Biomerieux, Marcy L’Etoile, France).^17^ We ran PCR for 45 cycles using the US CDC’s COVID-19 testing kit, which targets the N1 and N2 nucleocapsid proteins.^18^ We also ran PCR on each sample against the constitutive human enzyme RNAseP. The presence of RNAseP indicates that the NP swab made effective contact with the respiratory mucosa and that there were no PCR inhibitors.

We defined a COVID-19 test result as positive if the PCR cycle threshold (Ct) was <40 for both the N1 and N2 genes, if each assay demonstrated a logarithmic fluorescence amplification curve, had RNAseP detectable at Ct<40, and had positive and negative plate controls performing as anticipated.

However, Ct is a continuous variable, and the selection of any Ct cut point is arbitrary and hence controversial.^19^ We further note that most COVID-19 surveillance studies assess individuals who are acutely ill when viral loads, and hence PCR signals, are likely at peak intensity. By contrast, the population in this study were tested after they had died. At that point, the viral load and PCR signal intensity could have declined (or be undetectable), even if COVID-19 had set in motion a set of events culminating in death. Accordingly, we also report the number of decedents with COVID-19 detectable between Ct >40 to ≤ 45, though we have not included these in our summary analyses.

#### Genetic sequencing of selected samples

To characterize the distributions of novel COVID-19 variants, we conducted genetic sequencing to detect viral variants on the subset of samples with a Ct <30. Prior experience has shown that nucleic acid concentrations are rarely sufficient for sequencing above this threshold. For this, we tested samples both from surveillance Rounds 1 and 2, describing shifts in dominant COVID-19 variants in Lusaka over the cumulative period of surveillance.

Total RNA was re-extracted from vortexed NP swab specimens using the QIAamp^®^ Viral RNA Mini Kit (Qiagen, Hilden, Germany) as prescribed by the manufacturer. Complementary DNA was synthesized using LunaScript^®^ RT SuperMix Kit (New England Biolabs, Ipswich, Massachusetts, USA) according to the manufacturer’s protocol and multiplex PCR was conducted using custom primers, which were designed using Geneious software version 10.0.9, as used previously to sequence the first COVID-19 case in Zambia using the Sanger method.^20 21^ The multiplex PCR generated overlapping amplicons which were then sequenced on the MinION (Oxford Nanopore Technology, United Kingdom) platform. The samples were multiplexed using the Oxford Nanopore native barcoding expansion kits 1–12 and 13–24 in combination with the ligation sequencing kit 109 (Oxford Nanopore). The data generated through the MinION was processed using the standard ARTIC bioinformatic pipeline (https://artic.network/ncov-2019/ncov2019-bioinformatics-sop.html). After removal of sequencing primers, the consensus sequences were deposited in the GISAID database (www.gisaid.org/). Lineages were then determined for each genome using Pangolin v3.1.16 (https://pangolin.cog-uk.io/) and nextclade v0.13 (https://clades.nextstrain.org/).

### Statistical analysis

Given the uncertainties of future detection rates for COVID-19, our sampling was not based on a *priori* power assumptions. Rather, we sought to enroll as many decedents as our team could reasonably accommodate. Our statistical analysis was a straightforward comparison of proportions that did not require statistical modeling or imputation. COVID-19 status, based on PCR, was stratified by setting (community vs. facility deaths), by age groups, by calendar date, and by geography (city ward).

Where indicated, we have adjusted prevalence estimates by calculating weighted outputs to account for the different sampling ratios used during enrolment. We calculated weights so that the sum of the weighted population would sum to the sampled population: unweighted sample (1,118) = weighted sample (1,118). This allowed us to calculate the relative frequency of our unweighted sample in the four weighted groups: (1) facility deaths < 1 year, (2) all other facility deaths, (3) community deaths < 1 year of age, and (4) all other community deaths. We sampled the facility deaths < 1 year, all other facility deaths, and community deaths in infants < 1 year at a 1:1 ratio, and the community deaths ≥ 1 year at a 1:3 ratio. We then calculated the weighted sample by multiplying the inverse of the sampling ratio and calculated the relative frequency of the weighted sample. Setting the weighted sample size at the actual enrolled sample size of 1,118, yielded sample weights of 0.53 for the portion of the sample that we recruited as 1:1 and the sample weight of 1.60 for the portion of the sample that we recruited as 1:3. Hence, the ratio of these sample weights, adjusting for rounding, is 1.60/0.53 =∼3:1.

For analyses of COVID-19 deaths over time and place and for the genetic sequence data, we combined results from Rounds 1 and 2 of surveillance. Effectively this provides a time series analysis over a 1-year period minus the 3-month gap from October 2020-Jan 2021 defined by the end and start of adjacent funding cycles. Where appropriate, we reference our results from Round 1 to compare and contextualize results from Round 2. For all statistical analyses and most data manipulations, we used SAS software (Cary, North Carolina, USA).

For the geospatial analyses, we used ArcGIS software (ESRI, Redlands, California, USA). We pulled population size data from the Zambia Data Hub: https://zambia-open-data-nsdi-mlnr.hub.arcgis.com/, which is managed by the Government of Zambia through the Ministry of Lands and Natural Resources, and the Zambia Statistics Agency. We downloaded these ArcGIS layers to the level of Lusaka’s city wards. We then created a 2-dimensional heat map that indexed the total number of CV19+ deaths against total enrolled deaths across each of Lusaka’s main city wards.

## RESULTS

### Study overview and summary of PCR results

This second round of surveillance spanned January-June 2021. The newly enrolled cohort included 1,118 deceased individuals ranging in age from <1 to 102 years. Contemporaneously, 6,270 deaths were reported in the official Lusaka death registry. Thus, our sample represented 17.8% of all deaths in this period. The age distribution of the enrolled vs the total deceased population were similar, arguing to representativeness (**Supplementary Figure S1**).

PCR was successfully conducted on 100% of the NP swabs, and 1,116 had an RNAseP <40, indicating adequacy of sample collection. COVID-19 was detected among 29.3% (327/1,116) of decedents without weighting, and 32.0% (effective sample size = 358/1,116) after adjusting for weighting (**Table 1**). Thus, the overall prevalence of COVID-19 in Round 2 was roughly double what we observed in Round 1 (16%, 58/364). The median Ct values for the N1 and N2 targets were ∼33 in both cases (**Supplementary Figure S2)**. An additional 58 samples (45.8 weighted) had COVID-19 detectable at a Ct ≥40-45 but are not included in subsequent results.

**Table 1.** Postmortem COVID-19 PCR results, stratified by setting (community vs. facility)

| **Setting of death** | **Negative, CT≥40** | **Positive, CT<40** | **Totals** |
| --- | --- | --- | --- |
| **Community %**<br>**(n/N)** | 66%<br>(359/543) | 34%<br>(185/543) | 49%<br>(544/1,116) |
| **Facility %**<br>**(n/N)** | 75%<br>(431/573) | 25%<br>(142/573) | 51%<br>(574/1,116) |
| **Both settings combined**<br>**(unweighted)** | 71%<br>(789/1,116) | 29%<br>(327/1,116) | 100%<br>(1,116/1,116) |
| **Both settings combined**<br>**(weighted)** | 68%<br>(757/1,116) | 32%<br>(358/1,116) | 100%<br>(1,116/1,116) |

The weighted proportion of females among the COVID-19+ and COVID-19-groups was nearly identical (40.6%, 453/1,116). By contrast, the COVID-19+ decedents were significantly older than those without COVID-19 ((median 48 years (IQR 30-70 years) vs. 39 years (IQR 0-58 years), Wilcoxon Ranked Sum test *p*-value < 0.001 (**Table 2**)).

**Table 2.** Basic demographics of the postmortem cohort, stratified by COVID-19 status

|  | **Negative,**<br>**CT≥40** | **Positive,**<br>**CT<40** | **All Deaths** |
| --- | --- | --- | --- |
| **Parameter** | (n=791) | (n=327) | (n=1,118) |
| **Unweighted percent Females**<br>**(n/N)** | 42.2%<br>(334/791) | 41.0%<br>(134/327) | 41.9%<br>(468/1,116) |
| **Weighted percent female**<br>**(n/N)** | 41.0%<br>309/758 | 39.9%<br>144/358 | 40.6%<br>453/1,116 |
| **Unweighted median age at death,**<br>**years (IQR)** | 32.5<br>(0 - 54) | 44.0<br>(27 - 68) | 36.0<br>(1 - 58) |
| **Weighted median age at death (IQR)** | 39.0<br>(0-58) | 48<br>(30-70) | 41<br>(23-64) |

### Proportion of deaths and COVID-19+ deaths by facility vs. community setting

Within the 1,116/1,118 (99.8%) of total deaths with a valid test result, there were 573 facility deaths and 543 community deaths. While this implies that the ratio of facility to community deaths was similar, this does not account for the different enrollment ratios applied to each group. After weighting, 73% of all deaths occurred in the community vs. 27% at a facility, for a ratio of ∼three community deaths for each facility death.

COVID-19 was detected among 24.8% (142/573) of facility deaths and 34.0% (185/543) of community deaths. Considering the contribution of deaths by setting, among the 327 decedents with COVID-19, 56.6% (185/327) were community deaths and 43.4% (142/327) were facility deaths. After weighting, community deaths accounted for 79% and facility deaths for 21% of total COVID-19+ deaths. Stated another way, for each COVID-19+ death at a facility, ∼four COVID-19+ deaths occurred in the community.

### COVID-19 deaths by age group

During Round 1, the proportion of decedents with COVID-19 was similar across all age groups at, and 10% of all COVID-19+ deaths were in children ≤19 years.

Round 2 yielded a similar pattern with COVID-19+ deaths occurring across all age groups and with relatively similar proportions out of total deaths in each group. After weighting, adults aged >19 years comprised 86.2% (300/358) of COVID-19+ deaths and children 0 to ≤19 years for 14.9% (53/358) (**Table 3**). Overall, 78.1% (208/358, weighted) of COVID-19+ decedents were aged <60 years. Contrasting the death by age distributions for the COVID-19+ decedents relative to the age distributions for the total population from the burial registries, the COVID-19+ deaths were relatively underrepresented among children and relatively over represented in the very elderly. Outside of the age extremes, the age by death distributions for the two cohorts were similar (**Supplemental Figure S1**).

**Table 3.** Distribution of CV19 positive deaths by 20-year increments

| Age at death (years) | Ct>=40 (Neg) | Ct<40 (Pos) | Total | Ct>=40 (Neg) | Ct<40 (Pos) | Total |
| --- | --- | --- | --- | --- | --- | --- |
|  | Unweighted |  |  | Weighted |  |  |
| 0-19 | 308<br>82.8% | 64<br>17.2% | 372 | 205.6<br>79.4% | 53.3<br>20.6% | 261.0 |
| 20-39 | 159<br>66.5% | 80<br>33.5% | 239 | 184.8<br>67.1% | 90.5<br>32.9% | 275.3 |
| 40-59 | 171<br>73.4% | 62<br>26.6% | 233 | 190.1<br>74.8% | 63.9<br>25.2% | 253.9 |
| 60-79 | 108<br>58.1% | 78<br>41.9% | 186 | 120.3<br>55.9% | 94.8<br>44.1% | 215.1 |
| 80-99 | 42<br>52.5% | 38<br>47.5% | 80 | 55.4<br>52.0% | 51.1<br>48.0% | 106.5 |
| 100+ | 0<br>0.0% | 2<br>100.0% | 2 | 0<br>0.0% | 3.2<br>100.0% | 3.2 |
| Total | 788* | 324* | 1,112* | 757.7* | 358.4* | 1,112.1* |

\*4 had missing age data, of which 3 were CV19 positive and 1 was CV19 negative, for 789, 327 and 1,116, respectively
Row results are N (Top) and % (Bottom) in each cell.

### Antemortem testing for COVID-19

During Round 1, antemortem testing for COVID-19 was rarely done, occurring in only 10% of facility deaths and none of the majority of deaths that were in the community. In Round 2, testing rates had improved markedly among facility deaths. Within this group, 52.6% (302/574) were tested antemortem, of which 25 were reportedly positive (see **Supplementary Table S1**). Most testing at UTH relied on rapid antigen tests during this time. However, among the community deaths, we were still only able to document antemortem testing for 1.8% (10/544) of COVID-19+ decedents. Given that the burden of COVID-19 deaths in the community was four-times greater than at facilities, this indicates a substantial undercount of COVID-19 deaths.

### Causality

During Round 1, among those who had sufficient clinical data to allow an assessment of causality, nearly all were judged to have probably or possibly died from COVID-19. In Round 2, 89.3% (292/327) of deaths had sufficient data for adjudication, leaving 10.7% (45/327) that could not be adjudicated. Excluding the pediatric cases and those with insufficient data for analysis, we were able to adjudicate 219 deceased adults. Of these, 155/219 (70.8%) were considered ‘probably’ or ‘possibly’ due to COVID-19. When adjusted for weighting, this proportion rose slightly to 73.9% (181/244). Thus, a majority of evaluable adults positive for COVID-19 had a clinical syndrome implicating COVID-19 as the cause of death (see **Table 4**).

**Table 4.** Causal role of COVID-19 among PCR(+) deceased adults*, >19 years, unweighted and weighted cases

| **Role of COVID-19 in death of patient** | **Cases (N)** | **Percent** | **Weighted cases (N)** | **Percent** | **Cumulative total for cases that could be adjudicated** | **Cumulative percent (n/N) for cases that could be adjudicated** |
| --- | --- | --- | --- | --- | --- | --- |
| **Probably due to COVID-19** | 127 | 48.7% | 142.2 | 46.6% | 142 | 58.2%<br>(142/244) |
| **Possibly due to COVID-19** | 28 | 10.7% | 38.3 | 12.6% | 180 | 73.8%<br>(180/244) |
| **Probably not due to COVID-19** | 64 | 24.5% | 63.9 | 20.9% | 244 | 100%<br>(244/244) |
| **Insufficient data to adjudicate causality** | 42 | 16.1% | 60.7 | 19.9% | Not applicable | Not applicable |

\* Clinical presentations of children with COVID-19 (n= 63) were variable and often lacked typical symptoms of respiratory distress. Since we remain agnostic as to how COVID-19 should behave in children, we did not attempt to adjudicate these cases, but merely to describe these presentations (see **Figure 3** and **Supplementary Table S3**).

### Temporal shifts in COVID-19 prevalence, dominant variants, and geography

Given these changes between Round 1 and Round 2, we further explored the relationship between total deaths, COVID-19 prevalence, viral lineages, and geographic distribution in time series analyses. These results, combining Round 1 and 2 data, are presented in **Figure 2**.

**Figure 1.**
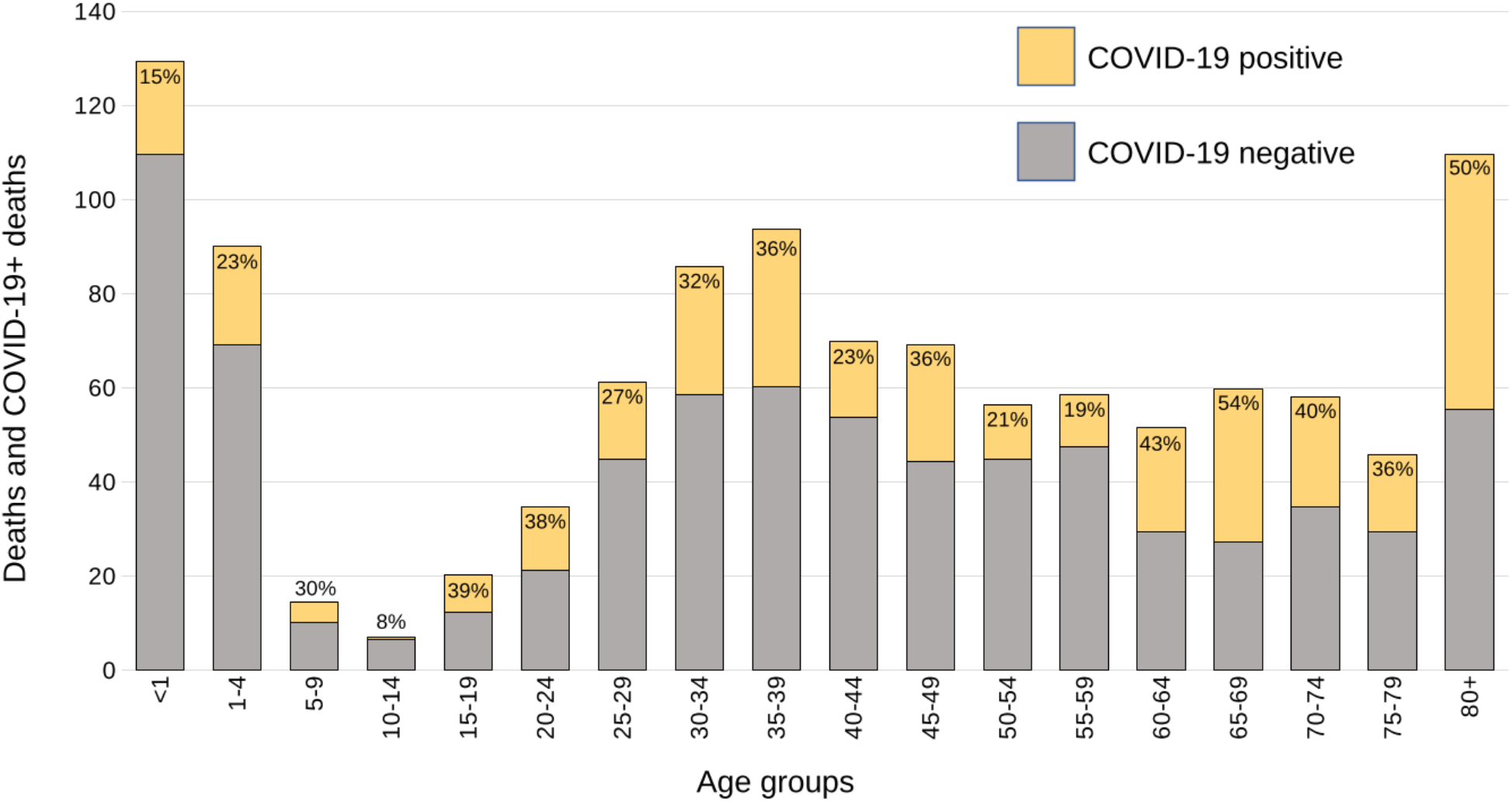
Enrolled decedents and COVID-19+ deaths by 5-year age strata Each stacked bar in the histogram shows the total number of enrolled decedents within each age stratum. The portion in yellow represents those who tested positive by PCR (Ct <40) for COVID-19 and those in grey those who tested negative (Ct ≥40). The % of positives is indicated at the top of each column. The totals have been adjusted to account for differing enrolment ratios.

**Figure 2.**
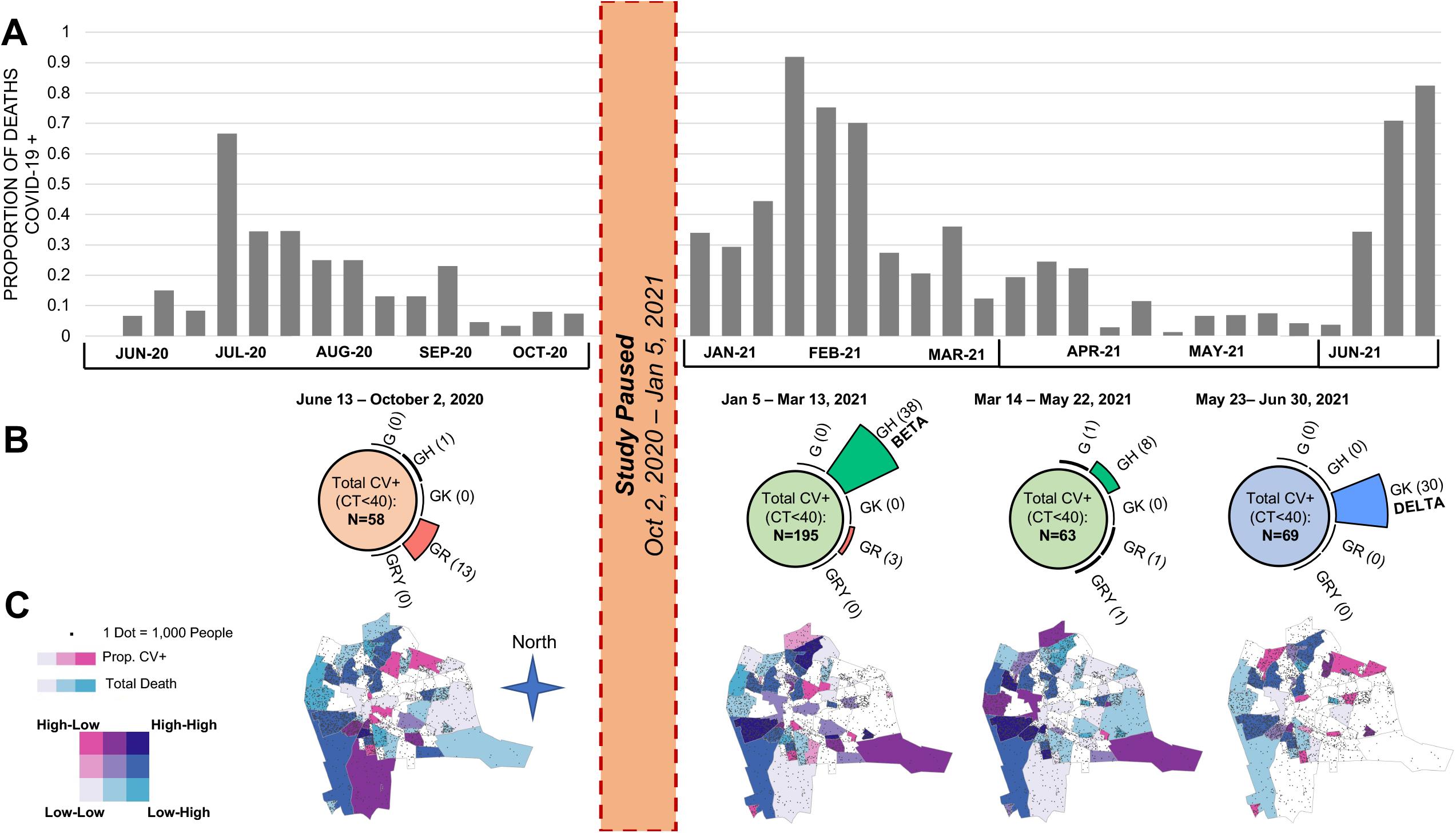
Proportion of COVID-19+ deaths and viral lineages/variants, by time and geography **Panel A**. The histograms represent the total number of COVID-19 positive deaths identified during the two rounds of surveillance. The gap from October 2020 to January 2021 represents the end and start of funding cycles. It is apparent that there were at least three waves of transmission in 2020-2021. Since our enrollment strategy shifted between Round 1 and Round 2, we have adjusted the proportions for the Round 2 data by weighting to account for the different enrolment ratios applied by age and setting. **Panel B**. Combing the samples corresponding to each of the three waves, and the period in between waves 2 and 3, we clustered the viral lineages as shown on the top half of the figure. As can be seen, wave 1 was dominated by lineage AE.1, wave 2 by the ‘Beta’ variant, and wave 3 by the ‘Delta’ variant. For these data, we present the results without adjustments for weighting. **Panel C**. Over these same time intervals, we summarize the distribution of cases indexed against total population within the major city wards of Lusaka. The colors correspond to a heat map defined by the union of ‘total deaths’ in pink and ‘COVID-19+’ deaths in blue, and therefore provide an indexed metric of COVID-19 deaths against all deaths. Those with darker shading indicate that COVID-19 deaths were disproportionately high relative to all deaths; those that are lightly shaded or white are where COVID-19 deaths were disproportionately low, relative to total deaths. As can be seen, the highest concentration of cases clustered in the peripheral areas of the city, which correspond to the poorest and most densely populated parts of the city. Whereas the wealthier sections, clustering in the middle of Lusaka, were relatively spared. Round 2 results have also been adjusted for enrollment ratios as in Panel A.

Several features are noteworthy. First, the data collected in Round 1 document what in hindsight was likely the first, and smallest, of three waves of COVID-19 to occur in Lusaka during our surveillance period (**Panel A**). Noting the gap in surveillance from October 2020 to January 2021, wave 1 occurred in June-October 2020; wave 2 in January-February 2021; and wave 3 in May-June 2021. Our surveillance ended in June 2021, precluding observations about when wave 3 peaked and receded.

Second, during waves 2 and 3, the proportion of COVID-19+ deaths grew explosively reaching 91.7% prevalence in January and 83.8% in June of 2021. Both represented a statistically significant increase over the average prevalence during Rounds 1 and 2 (ANOVA, P<0.001).

Third, each wave presented a distinct pattern in terms of prevalent viral lineages (**Panel B**). Combining Rounds 1 and 2 and selecting those with a Ct<30, we successfully sequenced 96 isolates. From these, during wave 1 lineage AE.1 was dominant. During wave 2 in Jan-Feb, the B.1.351 variant (the Beta variant) was dominant. This was supplanted by the abrupt emergence of the B1.617 variant (Delta) as wave 3, which accounted for 100% of all sequenced isolates during this period.

Concurrently, we mapped the distribution of COVID-19+ cases indexed against total deaths over time across Lusaka’s major city wards (**Panel C**). The areas shaded dark blue represent areas where COVID-19 deaths were disproportionately high relative to total deaths, while those in light pink, light blue, or white are areas where COVID-19 deaths were underrepresented relative to total deaths. Within this taxonomy, the heaviest burden of COVID-19 fell in Lusaka’s poorest and most densely populated areas that constitute the peripheral ring of peri-urban compounds to the city’s north, west, and south, and largely spared the more affluent neighborhoods that cluster in the center of the city.

### Clinical presentation of pediatric COVID-19 in this cohort

Recalling that infants <1 year were oversampled in our cohort, most deaths in children ≤19 y were in infants. With that caveat, **Supplementary Figure S3** summarizes the syndromic presentations for the COVID-19 positive deceased children <19 years, stratifying these by age groups and community/facility settings. Since we are describing these on a per individual basis, there was no need to weight these results.

The syndromic categorizations are not mutually exclusive. Clinical presentations appeared to vary by age and setting. Among infants who died at a facility, respiratory symptoms and fever predominated, whereas fever, respiratory and gastrointestinal symptoms occurred in similar proportions among those who died in the community. Among children 1-4 years, fever and gastrointestinal symptoms were more common than respiratory symptoms. Few deaths occurred in children aged 5-14 years (N=5). Among children 16-19 years (N=5, all from the community), the only reported symptoms were fever and respiratory distress, consistent with COVID-19’s presentation in adults. In both community and facility settings, there were several children with no reported symptoms, though this may simply reflect a lack of documentation. Acknowledging the sparse data from older children, these results are broadly consistent with our observations in Round 1 where we had noted the relative infrequency of respiratory symptoms and commonality of gastrointestinal symptoms among younger children. **Supplementary Table S3** provides a detailed line listing summarizing the clinical presentation of each CV19+ pediatric death by setting (facility deaths **Table S3a**, community deaths **Table S3b**)

## DISCUSSION

This second round of postmortem surveillance for COVID-19 reinforces and expands upon our previous observations. First, COVID-19 has had a severe impact in Lusaka with much loss of life. Second, such deaths were heavily concentrated in the community where testing for COVID-19 was essentially absent. Third, rather than being concentrated in the elderly, COVID-19+ deaths were distributed widely across the age spectrum, and most (∼80%) were in individuals aged <60 years. Fourth, COVID-19 was frequently identified among children. Among children <5 years, gastrointestinal complaints were common and respiratory symptoms comparatively uncommon. Lastly, the emergence of the Beta and Delta variants coincided with marked increases in the proportion of COVID-19+ deaths, reaching ∼90% prevalence during peak periods.

A consistent finding was the low proportion of decedents with COVID-19 who were diagnosed in life. We explain this as follows: 1) the majority of COVID-19+ deaths (∼80%) occurred in the community where, with few exceptions, testing was unavailable; 2) among the hospital deaths, only half were tested; and 3) most of the testing done at facilities relied on rapid antigen tests, which are far less sensitive than PCR.^22^ Putting these in sequence, from the 1,118 decedents tested in our study we identified COVID-19 in 327 (unweighted). Of these, only 25 had been identified antemortem at UTH, or <10% of the total. Since these hospital data feed into the national COVID-19 surveillance data, we conclude that the impact of COVID-19 has been substantially and systematically under-represented.

As in our prior analysis, the fatal burden of COVID-19 deaths was concentrated in Lusaka’s most densely populated wards where Lusaka’s poorest and most vulnerable citizens reside. Access to care is challenging in these communities, and we have described how delays in seeking care often contribute to infant deaths in the community.^23^ This provides a sad element to the global COVID-19 narrative that we suspect is underappreciated yet far from unusual, which is that COVID-19’s impact is not shared equally. Even within a poor country like Zambia, there is a gradient of impact that falls hardest on those with the least resources to protect themselves.

There are several important distinctions from our findings in Round 1. First, the numbers of COVID-19 deaths and the proportion of deceased individuals who tested positive for COVID-19 increased significantly.

Second, the proportion of these deaths that were probably or possibly judged as ‘due to COVID-19’ declined from ∼100% in Round 1 to ∼70 % in Round 2. Our data do not provide an explanation for this decline, but several hypotheses are plausible. One is that earlier population exposure to COVID-19 (following the first wave) might have provided increased resistance to the SARS-CoV2 virus, i.e., the change is explained by natural immunity from prior infection. COVID-19 vaccines were virtually unavailable during this period, so vaccine derived immunity cannot explain this difference. A second possibility is that the Beta and Delta variants dominated the second and third waves of transmission. Both variants exhibited increased transmissibility, which plausibly accounts for the high proportions of deaths with COVID-19 at the peak of each wave. As seen with the Omicron variant, a gain in transmissibility could yield a reciprocal loss of virulence, i.e., the change is explained by virology.^24 25^ However, recent epidemiologic and *in vitro* data suggest the opposite, that the Beta and particularly Delta variants were more, not less, pathogenic.^26-28^ Thus, the former hypothesis seems more plausible.

Third, the proportion of COVID-19 infections identified antemortem increased, but only among the facility deaths. No such improvements occurred in the community COVID-19 deaths, who constituted a four to one majority.

### Strengths and limitations

A strength was that our data were collected prospectively, capturing a wide spectrum of ages in community and facility settings and without prior knowledge of clinical presentation or antemortem testing results. And while all came from the UTH morgue, 80% of all deaths in the city transit this facility, making it highly representative. Additionally, we were able to combine multiple data elements on the same individual: clinical presentation, molecular testing, geography, and viral lineage, to provide an overall picture of the mortal impact of the COVID-19 pandemic in Lusaka over time.

Limitations include that there was a three-month gap in surveillance between Rounds 1 and 2, and that Round 2 ended prior to the resolution of Wave 3 caused by the Delta variant. Our adjudications were necessarily limited by the completeness of clinical data for the facility deaths and by the accuracy of recall from non-medical persons through the verbal autopsy for community deaths. That could lead to some misclassification that might affect causal inferences. Our data describe impact but cannot directly provide a case fatality rate since we lack concurrent incidence data. Lastly, while Round 2 data confirm the surprisingly high proportion of pediatric deaths with COVID-19 and again show that gastrointestinal symptoms are common in children <5 years, we are no closer to inferring causality. In our judgement, resolving this question requires a higher gold standard. The recently initiated Round 3 of our surveillance is incorporating histopathology and testing for COVID-19 in tissues to explore causality in children. Our data do not address the impact of COVID-19 on total mortality, but this question is being examined in an ongoing excess mortality analysis to be published separately.

## Conclusions

COVID-19 has had a devastating impact in Lusaka. Despite increased rates of antemortem testing for COVID-19, there has been little commensurate increase in antemortem testing for those who died in the community, who constituted the majority of all deaths. Overall, only about 10% of those who died with COVID-19 were identified in life. Considering recent reports demonstrating examples of underreporting of COVID-19 in other settings, including inferential methods based on excess mortality analyses,^9 29-34^ we believe that our results are typical rather than exceptional. If so, we again conclude that COVID-19’s impact across Africa has been vastly underestimated.

## Data Availability

All data produced in the present study are available upon reasonable request to the authors

## ACKNOWLEDGEMENTS

We wish to thank our team at the Bill & Melinda Gates Foundation for their support and encouragement: Prachi Vora, Padmini Srikanthia, and Keith Klugman.

## DECLARATIONS

### Dissemination of results

Round 2 data were presented to members of the Zambian Ministry of Health and the Zambian Medical Association in July 2021, and the final manuscript was shared with the Zambian National Health Research Authority (NHRA) prior to submission. The results have been presented to our team at the Bill Ȧ Melinda Gates Foundation and to representatives from the US CDC and the Swiss Tropical Medicine Institute. These data have been shared with a modeling team at the Imperial College London to support our ongoing excess mortality analyses. Data from Round 1 were published previously in the BMJ.

### Ethical approvals

Ethical oversight for ZPRIME and the COVID-19 expansion were provided by the institutional review boards at Boston University and the University of Zambia. Written informed consent was obtained from the deceased’s family members or representatives.

### Data sharing

All data from this analysis can be shared based on a reasonable request that includes a statement of the proposed study’s objectives and an analysis plan through a formal data sharing agreement between both parties. Similarly, we are willing to share biological samples based on any reasonable request and proposal and pending the completion of an appropriate materials transfer agreement authorized by the Government of Zambia.

### Community engagement

Due to the time constraints around the pandemic, we did not engage the community on this project.

### Data provenance

The lead authors (CJG and LM) affirm that the manuscript is an honest, accurate, and transparent account of the study being reported; that no important aspects of the study have been omitted; and that any discrepancies from the study as originally planned (and, if relevant, registered) have been explained.

### Manuscript contributions

**CJG** was the principle investigator, conceived the project, secured the funding, assisted with the protocol and led on writing the manuscript; **LM** was the Zambian PI, assisted with protocol and tool development, and oversaw the field team and study implementation; **WBM** was the study statistician, developed the analysis plan and conducted the analysis; **GK** was the technical lead for the PCR lab in Lusaka; **RP** was the project manager and a research fellow, developed the study proposal, oversaw study management, and contributed to key aspects of data analysis and presentation; **LE** was a research fellow and contributed to key aspects of data analysis and presentation; **RN, Chilufya Chikoti (CC#1**) and **BY** were the PCR technicians for our lab; **SC, LP, GM, DN** were the field data collection team under team leaders **BN** and **Charles Chimoga (CC#2, unrelated to CC#1**), who conducted grief counseling for the families, obtained consent, and collected all of the clinical data and the biological samples: **LF** supervised creation of the RedCAP data collection tools and oversaw data management, cleaning and retention; **RL** contributed to data interpretation and analysis; **JL** provided information technology support to the field team; **BK, JM, BM, MM** and **WM** conducted the genetic sequencing analyses under the supervision of **NS, DB** and **ES**; **ZM** was the PCR lab manager; **NP** was a research fellow who supported the analyses; and **DMT** was our senior co-investigator who contributed to data analysis and interpretation.

**Supplementary Figure S1**. Comparison of death by age distributions for the total deceased cohort, the enrolled cohort, and the COVID-19 positive enrolled cohort.

Data for the total death cohort were from Lusaka’s burial registry logs maintained by the Zambian Ministry of Health. From each entry, we extracted data of death, age, and sex. We then extracted those deaths that corresponded to the periods of surveillance to capture the age by death distribution for the total cohort (enrolled and unenrolled). The second two sets correspond to the total enrolled cohort (N=1,118 person) and the subset who were COVID-19+ (N=327).

**Supplementary Figure S2**. Distribution of cycle threshold values for the N1 and N2 PCR reactions

The median Ct results were 32 and 33, respectively for the N1 and N2 targets. Results for detections between >40 and 45 are noted in text but are not included in any of our summary results otherwise.

**Supplementary Figure S3**. Clinical presentations among COVID-19 positive children ≤19 years

For this analysis, we extracted the data about clinical presentation of each case from the medical charts (facility deaths) or verbal autopsy data (community deaths). We then clustered these syndromically into those that were ‘respiratory’ vs. ‘gastrointestinal’ vs. ‘other’. Since ‘fever’ could occur in any of these, or by itself, we summarized this separately. As can be seen, the syndromic presentation of COVID-19+ infants <1 is distinct from that in older children. In the infants, we see a high proportion with gastrointestinal complaints and a relative paucity with respiratory disease. In the older children, this pattern reverses, and in adolescents the pattern is consistent with that seen in adults, i.e., various combinations of respiratory symptoms, often accompanied by fever. Since this is an analysis by individual, there was no need to make weighting adjustments to account for enrollment ratios.

