## Supplementary tables and figures for "Sustained high prevalence of COVID-19 deaths from a systematic post-mortem study in Lusaka, Zambia: one year later"

**SUPPLEMENTARY DATA**

**Table S1**. Proportion of the deceased who had been tested antemortem, by setting

|  | Facility | Community | All deaths |
| --- | --- | --- | --- |
| Tested antemortem | 47.4% (272) | 1.8% (10) | 282 |
| Not tested antemortem | 52.6% (302) | 97.2% (534) | 836 |
| Total | 574 | 544 | 1,118 |
| **Note:** We have elected not to conduct a detailed analysis of the concordance between the off study antemortem (clinical care) and postmortem (research) testing. Our team had no role in how those off study samples were collected, what swabs were used, how staff were trained, what test kits were used over time, what quality control measures were used, and so forth. Hence, we have no provenance over the non-study data and do not vouch for their accuracy.  With that caveat, we identified 35 individuals for whom a positive test result was recorded or reported (25 facility and 10 community deaths). Obviously, this is far fewer than what we observed in our postmortem testing. For the reasons outlined above, we can only hypothesize explanations for this discrepancy. Yet, the simplest explanation is plausible: most of the clinical samples were evaluated using rapid antigen kits, which are far less sensitive than PCR.  Please also note that this Table does not account for the different enrollment ratios between the facility and community deaths, which will under-represent the community deaths by three-fold. | | | |

**Table S2.** Proportion of the deceased who tested positive for CV19 by study month

|  | **Ct>=40**  **(Neg)** | **Ct<40 (Pos)** | **Total** | **Ct>=40 (Neg)** | **Ct<40 (Pos)** | **Total** |
| --- | --- | --- | --- | --- | --- | --- |
| **Month** | **Unweighted** |  |  | **Weighted** |  |  |
| January | 77 47.8% | 84 52.2% | 161 | 108.1 49.4% | 110.8 50.6% | 218.8 |
| February | 118 57.0% | 89 43.0% | 207 | 118.2 57.1% | 88.9 42.9% | 207.2 |
| March | 189 77.5% | 55 22.5% | 244 | 173.1 76.7% | 52.7 23.4% | 225.8 |
| April | 216 91.5% | 20 8.5% | 236 | 176.8 92.2% | 14.9 7.8% | 191.7 |
| May | 154 89.0% | 19 11.0% | 173 | 154.4 92.1% | 13.3 7.9% | 167.7 |
| June | 37 38.1% | 60 61.9% | 97 | 29.3 27.4% | 77.7 72.6% | 107.0 |
| Total | 791  70.8% | 327  29.2% | 1,118 | 759.9  68.0% | 358.4  32.0% | 1,118.3  100.0% |

Row results are N (Top) and % (Bottom) in each cell.

**Table S3.** Clinical presentations of pediatric CV19 positive deaths

| **Table S3a.** Clinical presentation of COVID-19+ facility deaths in children aged 0-19 years | | | | | |
| --- | --- | --- | --- | --- | --- |
| **No.** | **Age**  **in years** | **Respiratory**  **Symptoms** | **Gastrointestinal Symptoms** | **Fever** | **Cause of Death*** |
| 1 | <1 | difficulty breathing | diarrhea | X | sepsis |
| 2 | <1 | cough |  | X | pneumonia |
| 3 | <1 | cough, difficulty breathing, shortness of breath | diarrhea, vomiting | X | pneumonia |
| 4 | <1 | difficulty breathing |  |  | sepsis |
| 5 | <1 | cough, difficulty breathing |  | X | COVID-19 pneumonia |
| 6 | <1 | difficulty breathing |  |  | sepsis |
| 7 | <1 | difficulty breathing |  |  | pneumonia |
| 8 | <1 | difficulty breathing |  |  | sepsis |
| 9 | <1 | runny nose |  |  | sepsis |
| 10 | <1 | difficulty breathing |  |  | sepsis |
| 11 | <1 | cough | vomiting |  | sepsis |
| 12 | <1 | difficulty breathing |  | X | sepsis |
| 13 | <1 |  | vomiting | X | sepsis |
| 14 | <1 |  |  | X | sepsis |
| 15 | <1 |  |  | X | sepsis |
| 16 | <1 |  |  |  | sepsis |
| 17 | <1 | difficulty breathing, shortness of breath |  | X | sepsis |
| 18 | <1 |  |  | X | sepsis |
| 19 | <1 |  |  | X | sepsis |
| 20 | <1 | cough, difficulty breathing |  | X | sepsis |
| 21 | <1 | difficulty breathing |  |  | sepsis |
| 22 | <1 |  |  |  | hypothermia |
| 23 | <1 |  |  | X | sepsis |
| 24 | <1 | difficulty breathing |  |  | pneumonia |
| 25 | <1 | cough, difficulty breathing |  | X | pneumonia |
| 26 | 1-5 | shortness of breath |  | X | sepsis |
| 27 | 1-5 |  | diarrhea, vomiting |  | diarrhea |
| 28 | 1-5 |  |  |  | dehydration |
| 29 | 1-5 |  | diarrhea | X | malnutrition |
| 30 | 1-5 |  | diarrhea | X | sepsis |
| 31 | 1-5 |  |  | X | respiratory failure |
| 32 | 6-10 |  |  |  | diarrhea |
| 33 | 6-10 | difficulty breathing |  |  | sepsis |
| 34 | 11-15 |  |  |  | intracranial abscess |

* Cause of death from the official death certificate completed by the UTH medical examiner.

| **Table S3b**. Clinical presentation of COVID-19+ community deaths in children aged 0-19 years | | | | |
| --- | --- | --- | --- | --- |
| **No.** | **Age**  **in years** | **Respiratory**  **Symptoms** | **Gastrointestinal**  **Symptoms** | **Fever** |
| 1 | <1 | difficulty breathing |  |  |
| 2 | <1 | cough |  | X |
| 3 | <1 | cough, difficulty breathing | vomiting |  |
| 4 | <1 |  | vomiting |  |
| 5 | <1 |  |  | X |
| 6 | <1 |  | diarrhea, vomiting | X |
| 7 | <1 | cough, difficulty breathing, fast breathing, shortness of breath |  | X |
| 8 | <1 |  |  |  |
| 9 | <1 |  |  |  |
| 10 | <1 | cough, fast breathing |  | X |
| 11 | <1 |  | diarrhea, vomiting | X |
| 12 | <1 | cough, difficulty breathing | diarrhea | X |
| 13 | 1-5 | cough |  | X |
| 14 | 1-5 |  | diarrhea, vomiting |  |
| 15 | 1-5 |  | diarrhea, vomiting | X |
| 16 | 1-5 |  | diarrhea |  |
| 17 | 1-5 |  | diarrhea, vomiting | X |
| 18 | 1-5 |  |  |  |
| 19 | 1-5 |  |  | X |
| 20 | 1-5 |  | diarrhea |  |
| 21 | 1-5 | cough | diarrhea |  |
| 22 | 1-5 |  |  |  |
| 23 | 1-5 | cough, difficulty breathing |  |  |
| 24 | 6-10 |  |  |  |
| 25 | 6-10 |  | diarrhea |  |
| 26 | 16-19 | difficulty breathing |  |  |
| 27 | 16-19 | cough, difficulty breathing |  |  |
| 28 | 16-19 |  |  | X |
| 29 | 16-19 |  |  |  |
| 30 | 16-19 |  |  |  |

We do not include a cause of death for the community deaths. A death certificate is still issued for each case, but these are not based on information from medical providers and are not considered to be accurate.

**Figure S1.** Comparison of death by age distributions for the total deceased cohort, the enrolled cohort, and the CV19 positive enrolled cohort.

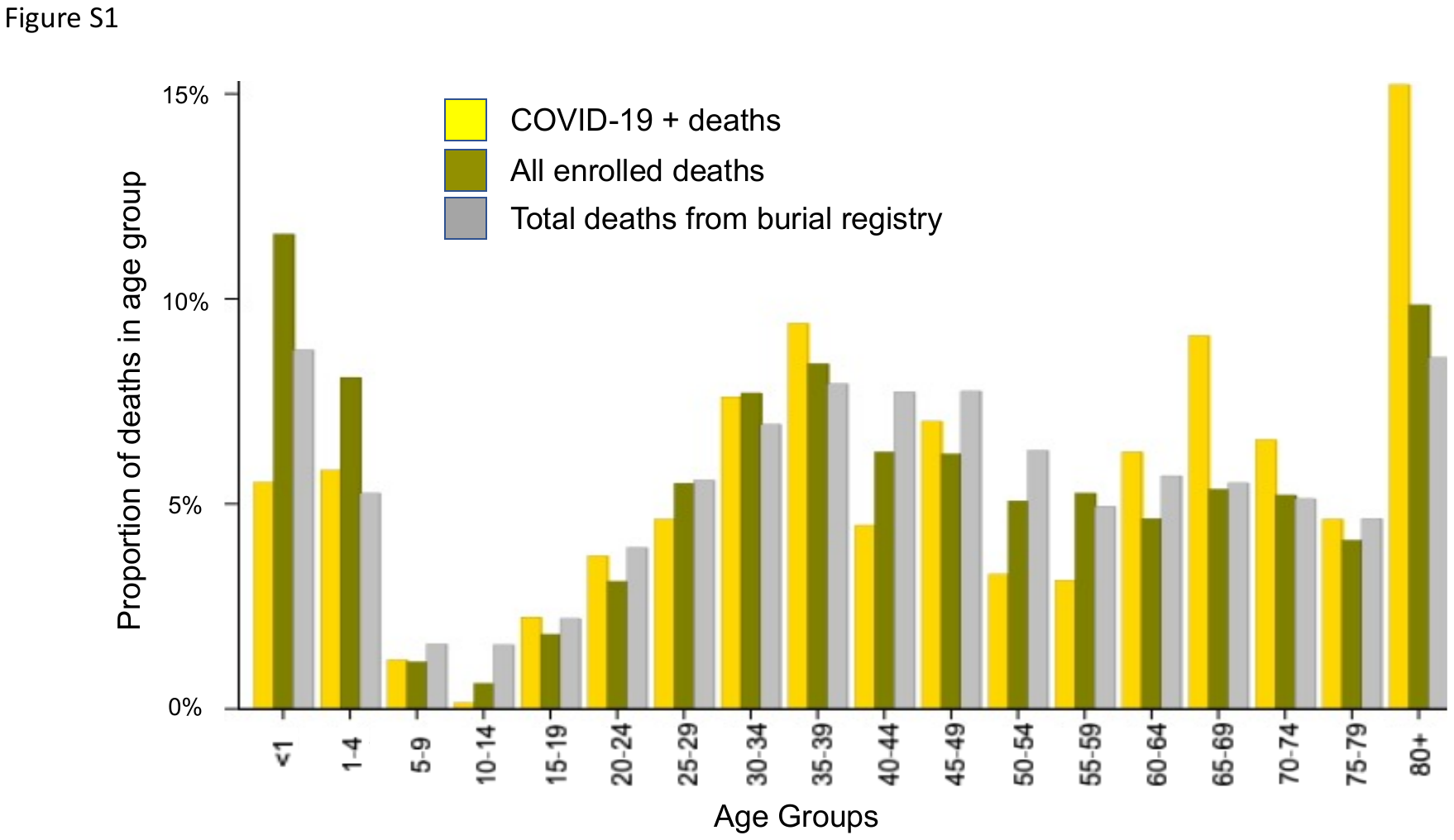

**Figure S2.** Distribution of cycle threshold values for PCR results targeting the N1 and N2 nucleocapsid proteins.

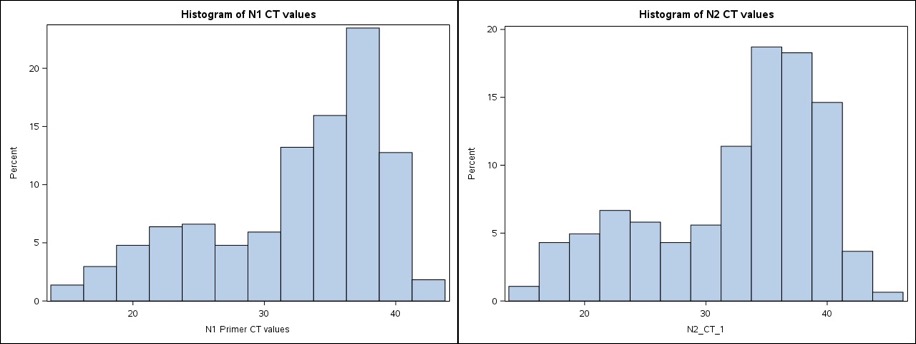

**Supplementary Figure S3**

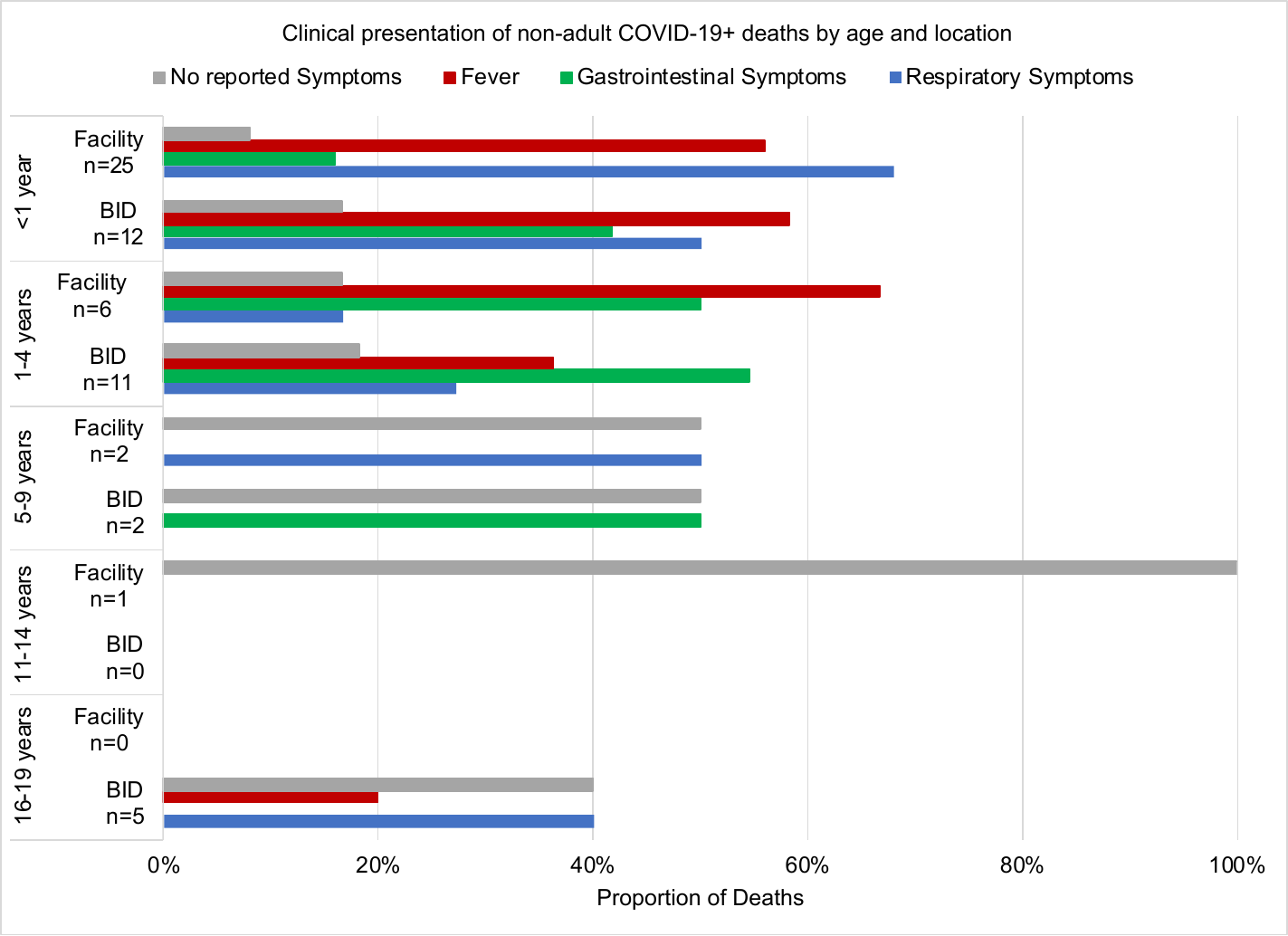
